## Supplemental Table 1 for "A Prospective Observational Study to Investigate Performance of a Chest X-ray Artificial Intelligence Diagnostic Support Tool Across 12 U.S. Hospitals"

**Supplemental Table 1:** Pre-implementation Temporal and Publically Available Validation

|  |  | | Temporal Validation | | | |  | Publically Available Validation | |
| --- | --- | --- | --- | --- | --- | --- | --- | --- | --- |
|  | All Patients | | | “Severe” Disease | | “Moderate” Disease | |  |  |
| Ratio^1^ | AUROC | AUPRC | | AUROC | AUPRC | AUROC | AUPRC | AUROC | AUPRC |
| 1 | 0.800 | 0.842 | | 0.834 | 0.871 | 0.704 | 0.742 | 0.960 | 0.975 |
| 2 | 0.801 | 0.759 | | 0.834 | 0.799 | 0.706 | 0.619 | 0.959 | 0.963 |
| 5 | 0.803 | 0.631 | | 0.834 | 0.688 | 0.705 | 0.440 | 0.961 | 0.949 |
| 12 | 0.801 | 0.497 | | 0.837 | 0.562 | 0.706 | 0.289 | 0.956 | 0.927 |
| 20 | 0.797 | 0.412 | | 0.829 | 0.483 | 0.705 | 0.218 | 0.965 | 0.929 |

^1^ Ratio between positive cases and negative case.
