## Supplemental Table 2 for "A Prospective Observational Study to Investigate Performance of a Chest X-ray Artificial Intelligence Diagnostic Support Tool Across 12 U.S. Hospitals"

**Supplemental Table 2:** External Validation

| **Institution** | **N** | **Prevalence** | **AUROC** | **Specificity*** | **Sensitivity*** | **PPV*** | **NPV*** |
| --- | --- | --- | --- | --- | --- | --- | --- |
| Indiana | 10,002 | 30% | 0.76 | 0.783 | 0.731 | 0.861 | 0.612 |
| Emory | 2,002 | 50% | 0.72 | 0.67 | 0.67 | 0.68 | 0.66 |

**Legend:**

Indiana University: Using the following thresholds for unlikely (score <= 0.03), indeterminate (score >0.03 – <0.13, *25% of images*) and likely (score >= 0.13) specificity, sensitivity, PPV, and NPV were obtained.

Emory University: Using the following thresholds for unlikely (score <= 0.02), indeterminate (score > 0.02 - <= 0.05), and likely (score > 0.05) specificity, sensitivity, PPV, and NPV were obtained.
