## Supplemental Table 3 for "A Prospective Observational Study to Investigate Performance of a Chest X-ray Artificial Intelligence Diagnostic Support Tool Across 12 U.S. Hospitals"

**Supplemental Table 3:** Real-time investigation of COVID-19 Diagnostic Algorithm performance

|  | **N** | **AUROC** | **95% CI** | **Prevalence** |
| --- | --- | --- | --- | --- |
| **Week 1** | 683 | 0.702 | 0.65-0.75 | 20.4% |
| **Weeks 8-19** | 5335 | 0.696 | 0.66-0.73 | 4.8% |
