## Supplementary figures and images for "A Prospective Observational Study to Investigate Performance of a Chest X-ray Artificial Intelligence Diagnostic Support Tool Across 12 U.S. Hospitals"

### Supplemental Figure 1

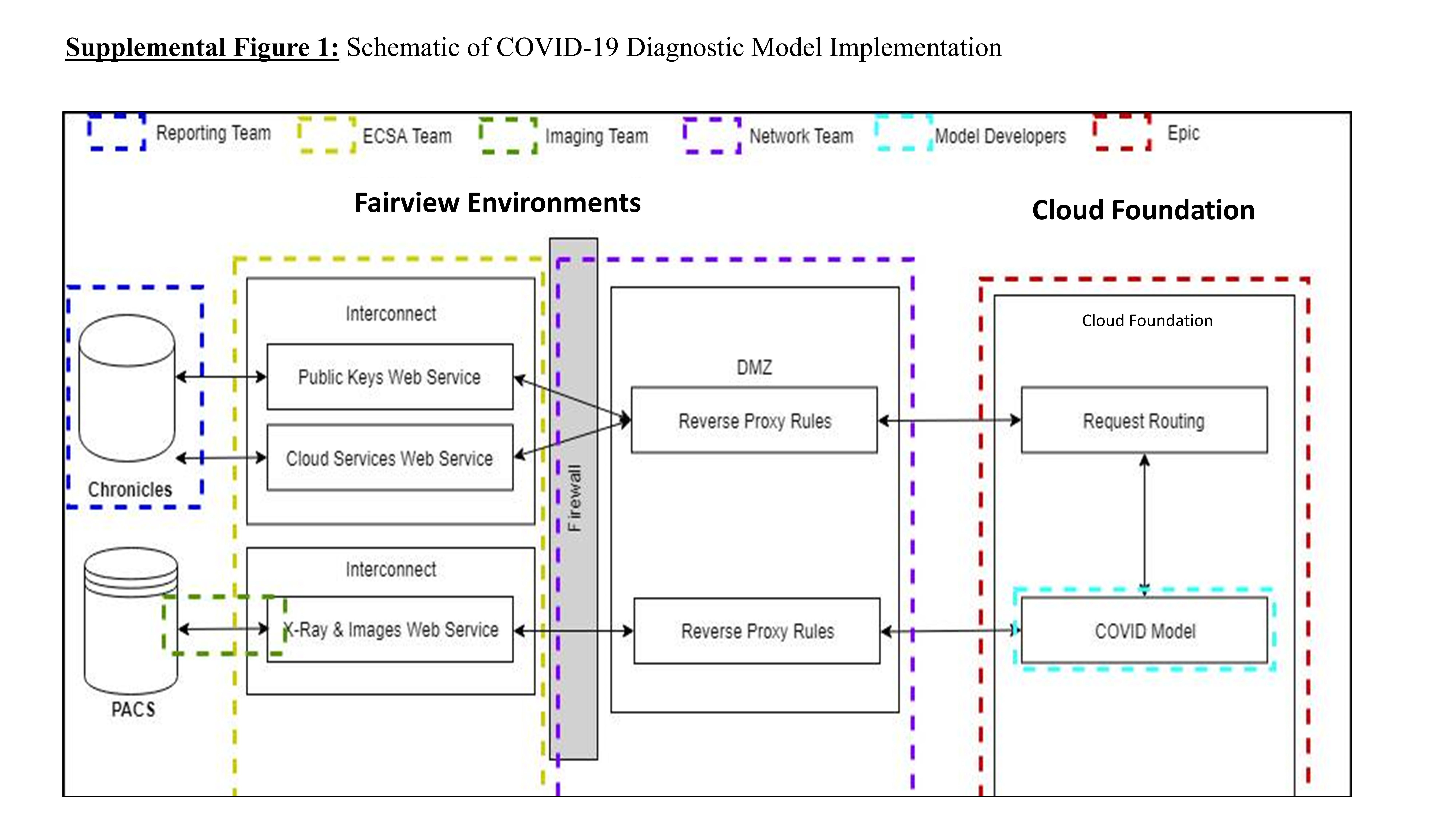

### Supplemental Figure 2

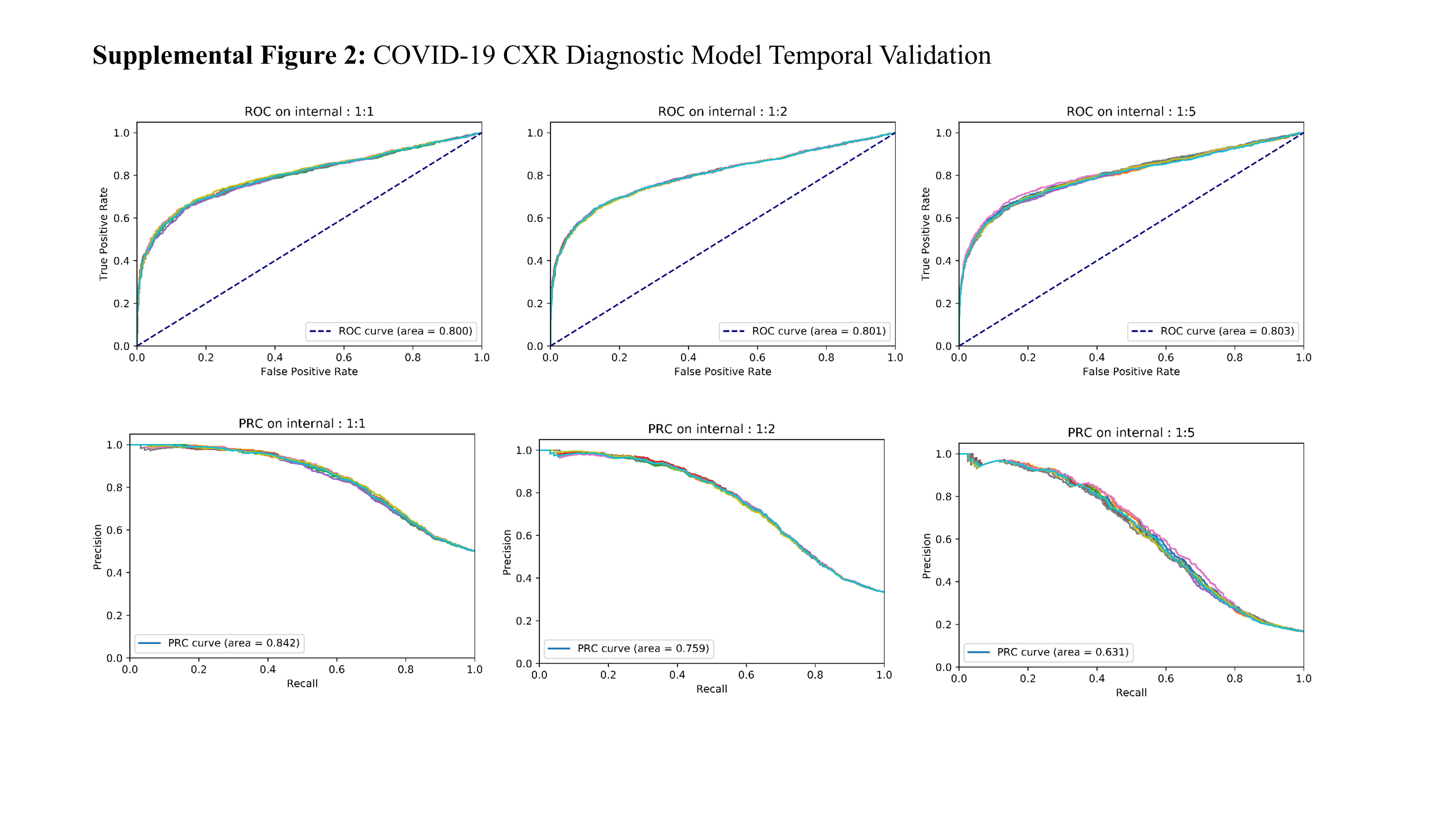

### Supplemental Figure 3

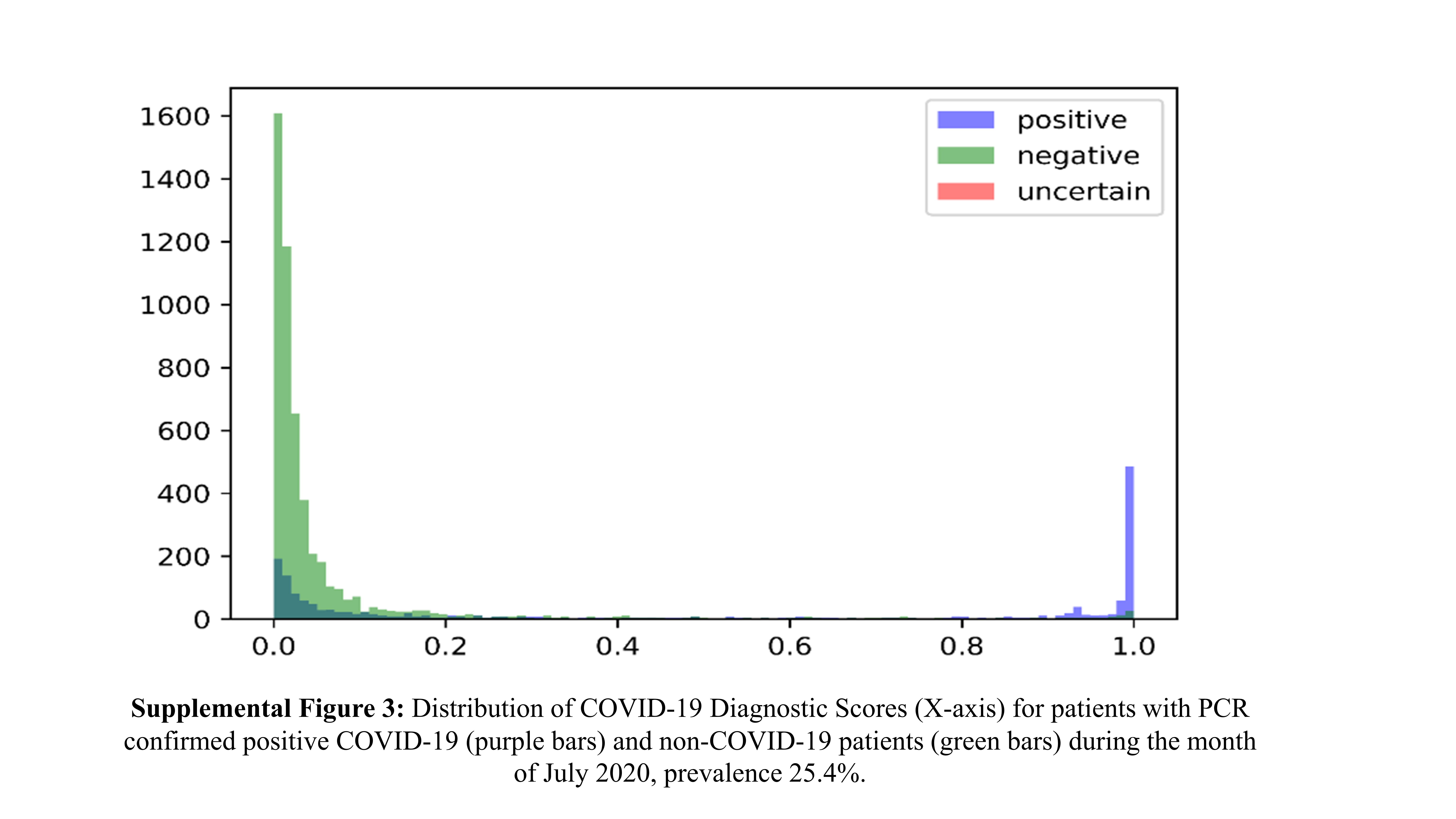

### Supplemental Figure 4

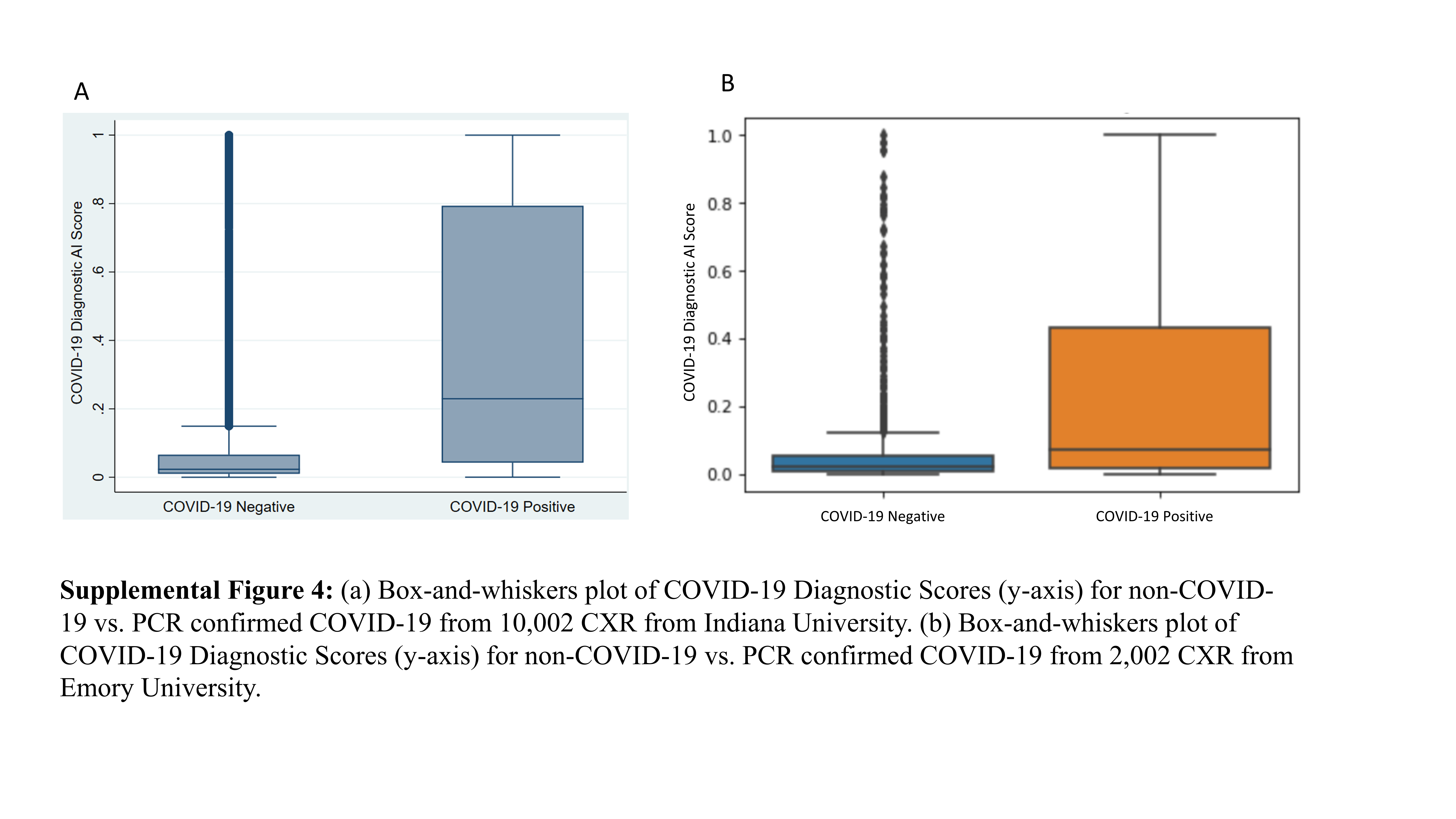
